# A novel strategy for the detection of SARS-CoV-2 variants based on multiplex PCR-MALDI-TOF MS

**DOI:** 10.1101/2021.06.08.21258523

**Authors:** Fei Zhao, Jianzhong Zhang, Xuemei Wang, Xuexin Hou, Tian Qin, Fanliang Meng, Xiaona Xu, Tianyi Li, Haijian Zhou, Biao Kan, Jinxing Lu, Di Xiao

**Author notes:** **Corresponding author** Di Xiao, Professor, Ph.D, 155 Changbai Road, Changping District, China National Institute for Communicable Disease Control and Prevention, Chinese Center for Disease Control and Prevention, State Key Laboratory of Infectious Disease Prevention and Control. Beijing 102206, China. **Competing interests** The authors declare that they have no competing interests.

## Abstract

**Background:** The second wave of coronavirus disease 2019 (COVID-19) has been incessantly causing catastrophe worldwide, and the emergence of severe acute respiratory syndrome coronavirus 2 (SARS-CoV-2) variants causes further uncertainty regarding epidemic risk. Here, a novel strategy for the detection of SARS-CoV-2 variants using multiplex PCR coupled with MALDI-TOF MS was developed.

**Methods:** Plasmids carrying gene sequences containing 9 mutation types in 7 mutated sites (HV6970del, N501Y, K417N, P681H, D614G, E484K, L452R, E484Q and P681R) in the receptor-binding domain of the spike protein of SARS-CoV-2 variants were synthesized. Using the nucleic acid sequence of SARS-CoV-2 nonvariant and a synthetic SARS-CoV-2-variant-carrying plasmid, a MALDI-TOF MS method based on the single-base mass probe extension of multiplex PCR amplification products was established to detect the above nine mutation types. The detection limit of this method was determined via the concentration gradient method. Twenty-one respiratory tract pathogens (9 bacteria, 11 respiratory viruses) and pharyngeal swab nucleic acid samples from healthy people were selected for specific validation. Sixteen samples from COVID-19 patients were used to verify the accuracy of this method.

**Results:** The 9 mutation types could be detected simultaneously by triple PCR amplification coupled with MALDI-TOF MS. SARS-CoV-2 and all six variants (B.1.1.7, B.1.351, B.1.429, B.1.526, P.1 and B.1.617) could be identified. The detection limit for all 9 sites was 1.5×10^3^ copies. The specificity of this method was 100%, and the accuracy of real-time PCR CT values less than 30 among positive samples was 100%. This method is open and extensible, and can be used in a high-throughput manner, easily allowing the addition of new mutation sites as needed to identify and track new SARS-CoV-2 variants as they emerge.

**Conclusions:** Multiplex PCR-MALDI-TOF MS provides a new detection option with practical application value for SARS-CoV-2 and its variant infection.

**Key point:** An all-in-one SARS-CoV-2 variant identification method based on a multiplex PCR-MALDI-TOF MS system was developed. All of the SARS-CoV-2 variants can be identified based on 9 types of 7 mutated sites of RBD of spike protein using this method.

## Introduction

Severe acute respiratory syndrome coronavirus 2 (SARS-CoV-2) has spread worldwide since the end of 2019. The second wave of COVID-19 has been incessantly causing catastrophe worldwide. The emergence of SARS-CoV-2 variants is the key factor in the second wave of the COVID-19 pandemic. To date, there are six different SARS-CoV-2 variants worldwide, including the United Kingdom (UK) variant (B.1.1.7), South African mutant (B.1.351), California mutant (B.1.429), New York mutant (B.1.526), Brazilian mutant (P.1) and recent India mutant (B.1.617) ^1, 2^. These variants have important mutations in the receptor binding domain (RBD) of the spike protein of SARS-CoV-2 that increase the transmission efficiency of the virus, increase the severity of the disease, enhance the immune escape ability of the virus, and reduce the immune effect of the current vaccines ^3-6^.

Currently, the mutation and evolution of SARS-CoV-2 are mainly detected by whole-genome sequencing (WGS) ^7-11^. Although genome sequencing is the gold standard for identifying SARS-CoV-2 variants, routine genomic testing is expensive and difficult to perform in real time and is not available in many areas due to lack of resources and expertise. qPCR ^12^, RT-PCR ^13^, and CRISPR-Cas13a -based transcription amplification were general used for the single-site detection of SARS-CoV-2 variants^14^. Therefore, the rapid, accurate, economic and multisite detection method of SARS-CoV-2 variant identification is an urgent technical system for SARS-CoV-2 infection prevention and control worldwide.

Matrix-assisted laser desorption/ionization time-of-flight mass spectrometry (MALDI-TOF MS) has been utilized for detecting SNPs in recent decades. Multiplex PCR was used to amplify the genes containing the targets of SNPs. Subsequently, an extension mass probe was utilized for the extension of SNP sites. Finally, MALDI-TOF MS was performed to identify the mass-to-charge ratio (m/z) of extended mass probes. In recent years, MALDI-TOF MS has been commonly used in clinical hospitals, the Center for Disease Control and Prevention, and research institutes in many countries for microbiological detection and analysis ^15-17^. To date, multiple SNP detection and diagnosis of various viral infections based on MALDI-TOF MS has been frequently conducted, and serves as an effective alternative to conventional techniques ^18-20^. Our team has also completed a multisite SNP genotyping and macrolide susceptibility gene method for *Mycoplasma pneumoniae* using multiplex PCR coupled with PCR-MALDI-TOF MS. SNP analysis based on MALDI-TOF MS features high-throughput, rapid detection and simultaneous detection of multiple targets^21^. Therefore, it is one of the most promising biomolecular techniques. In this study, 9 mutation types of 7 mutation sites in the RBD of the spike protein were detected simultaneously by using multiplex PCR-MALDI-TOF MS technology for the identification all current SARS-CoV-2 variants.

## Methods

### Specimens and nucleic acids of SARS-CoV-2

The nucleic acid of the SARS-CoV-2 clinical isolate (nonvariant) was obtained from the Institute of National Institute for Communicable Disease Control and Prevention, Chinese Center for Disease Control and Prevention (ICDC). The S gene mutation plasmids (plasmid 1 containing HV69-70del, K417N, E484K, N501Y, D614G, P681H mutation, plasmid 2 containing L452R, E484Q, P681R mutation) of SARS-CoV-2 variants (UK, South Africa, New York, California, Brazilian and India) were synthesized by Sangon Biotech (Shanghai, China). The nucleic acids of 21 common respiratory tract pathogens, including *Mycoplasma pneumoniae* (ATCC 29342), *Escherichia coli* (ATCC 11229), *Streptococcus pneumoniae* (ATCC 49619), *Staphylococcus aureus* (ATCC 29213), *Legionella pneumophila* (clinical isolate), *Mycobacterium tuberculosis* (clinical isolate), *Pseudomonas aeruginosa* (clinical isolate), *Hemophilus influenzae* (clinical isolate), *Neisseria meningitides* (clinical isolate), influenza A, influenza B, parainfluenza I, II, III and IV, adenovirus, rhinovirus, coronavirus 229E, OC43 and NL63, respiratory syncytial virus and normal human throat swab specimen were selected for specific validation of this method. The nucleic acids of 16 COVID-19 clinical samples (kept in ICDC) were used to verify the accuracy of this method. All the specimens collected from COVID-19 patients were collected with complete informed consent procedures. This study was approved by the ethics committee of ICDC.

### Design of amplification primer and extension probe for the variant sites

Three sets of PCR primers were designed to amplify the related fragments of the nine mutation types (Table 1). The S-F1/R1 amplification product contained one mutation type of HV69-70del, the S-F2/R2 amplification product contained five mutation types of K417N, E484K, E484Q, N501Y and L452R, and the S-F3/R3 amplification product contained three mutation types of D614G, P681H, and P681R. The length of primers ranged from 19 bp to 22 bp, and the molecular weight of each primer was more than 9 kDa (beyond the mass range of the mass extension probe) by adding a 10 bp fixed sequence (acgttggatg) to the 5’ end of each primer. MALDI-TOF MS quality difference probes for mutation sites were designed by using the IntelliBio genetic locus analysis software (V2.0, IntelliBio, China) (Table 1). The molecular weight of the mass probe was required to be 4 k-9 kDa and the length was 17-28 bp, and a minimal difference between the probe molecular weights was set for 20Da.

**Table 1.**
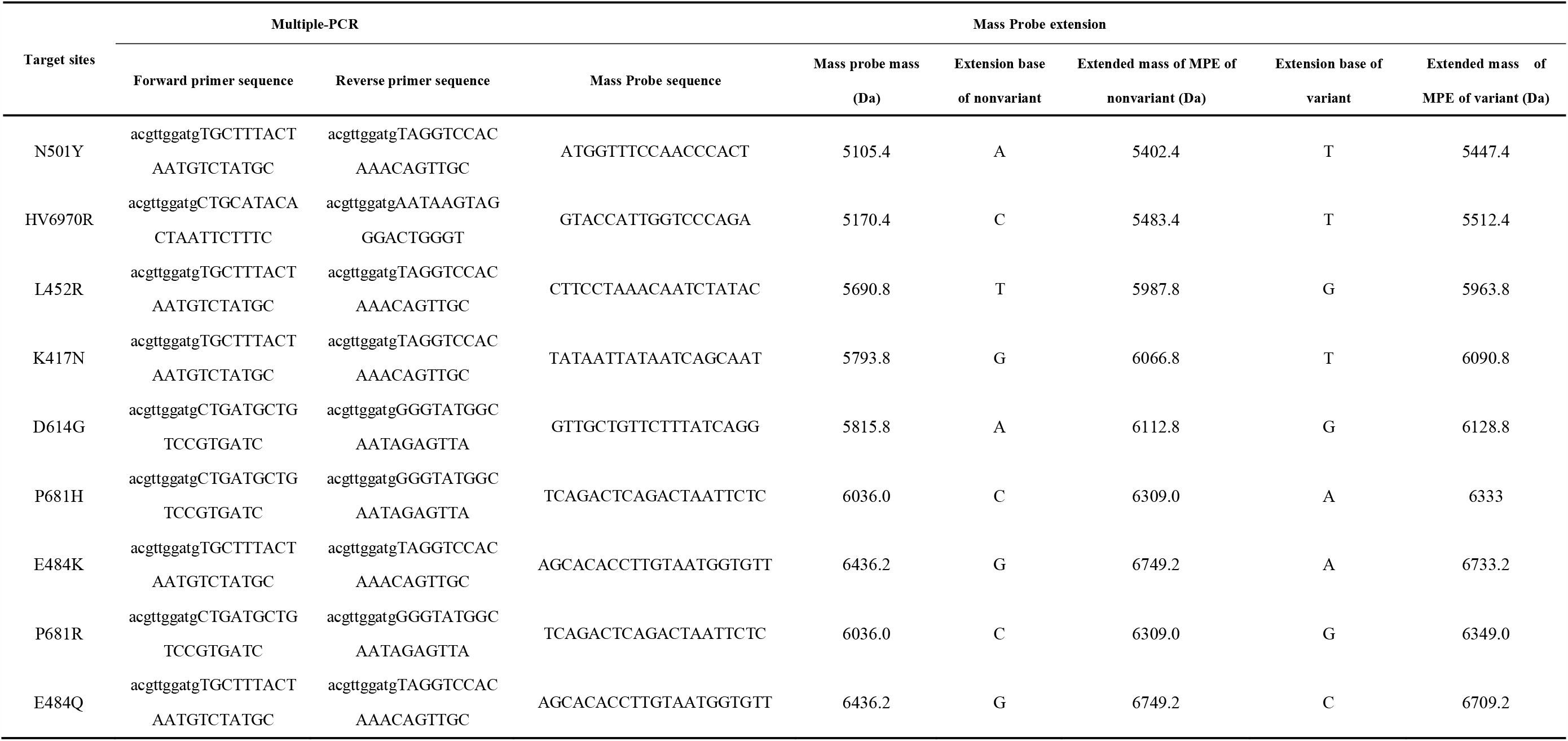
Primer sequences for SARS-CoV-2 variant target site amplification and MPE probes for target site detection.

### Establishment of PCR-MALDI-TOF MS method

SARS-CoV-2 nonvariants and S gene mutant plasmids of SARS-CoV-2 variants were used as templates to construct the analysis method. Nucleic acid-free water served as the blank control. The specific steps were as follows: (i) Multiplex PCR amplification: the mutated gene fragment was amplified by an AgPath-ID One-step RT-PCR kit; (ii) SAP digestion: PCR products were treated with shrimp alkaline phosphatase (SAP) to eliminate the free dNTPs; (iii) Mass probe extension (MPE): the purified PCR products were added with mixed MPE probes for single base extension, and the mutation sites were detected; (v) Desalination and purification: add an appropriate amount of resin to the extension product, turn over and mix for 30 min, centrifuge and detect the supernatant by MALDI-TOF MS.

### SNP identification and data analysis by MALDI-TOF MS

3-Hydroxypyridine-2-carboxylic acid (3-HPA, 0.9 μL) was dropped at the center of the sample target. After drying, the matrix was covered with 0.3 μL purified supernatant. Then, the samples were subjected to testing after crystallization. The data were acquired from the QuanTOF I system (Intelligene Biosystems, Qingdao, China). The parameters and data anslysis were previously described ^21^.

### Determination of the detection limit and specificity

The lowest detection limit (LDL) of PCR-MALDI-TOF MS was determined by using RNA of SARS-CoV-2 nonvariants and the mutant DNA plasmids of the S gene. The original concentration of nucleic acids was quantified (Qubit 3.0) and diluted in 7 concentration gradients. The specificity of this method was validated by 21 kinds of nucleic acids of the other respiratory pathogens (9 bacteria, 11 respiratory viruses) and human throat swab nucleic acid extraction.

### Nucleic acid validation of COVID-19 patients using multiplex PCR-MALDI-TOF MS

The nucleic acid of sputum and bronchoalveolar lavage fluid specimens of 16 COVID-19 patients was used to test the accuracy of this method. These specimens were also detected with a 2019-nCoV RT real-time PCR Detection Kit (Da An Gene Co., Ltd). All nucleic acid specimens were sequenced to confirm that there was no target mutation site detected in this study.

## Results

### Multiplex PCR-MALDI-TOF MS-based method establishment and optimization

The nine targets were amplified by triple PCR. The m/z of MPE original peaks at 9 mutation types (HV69-70del, N501Y, K417N, P681H, D614G, E484K, L452R, E484Q, and P681R) were 5170±3, 5105±3, 5794±3, 6036±3, 5816±3, 6436±3, 5691±3, 6436±3, and 6036±3, respectively (Mass error less than 500 ppm, the same below). The m/z of MPE extension peaks of these 9 mutation types of SARS-CoV-2 nonvariants were 5483±3, 5402±3, 6067±3, 6309±3, 6113±3, 6749±3, 5988±3, 6749±3 and 6309±3, and the extension bases were C, A, G, C, A, G, G, G, and C. The m/z of MPE extension peaks of these 9 mutation types of the S gene mutated plasmid of SARS-CoV-2 variants were 5512±3, 5447±3, 6091±3, 6333±3, 6129±3, 6733±3, 5964±3, 6709±3, and 6349±3, respectively, and the extended bases were A, T, T, A, G, A, G, C and G (Fig. 1). All mutation types were identified correctly using multiplex PCR-MALDI-TOF MS methods. After optimization, the final concentrations of 7 MPEs were 7.0 μM (HV69-70del), 8.09 μM (K417N), 9.09 μM (E484K/Q), 6.87 μM (N501Y), 8.12 μM (D614G), 8.48 μM (P681H/R), and 7.91 μM (L452R).

**FIG 1.**
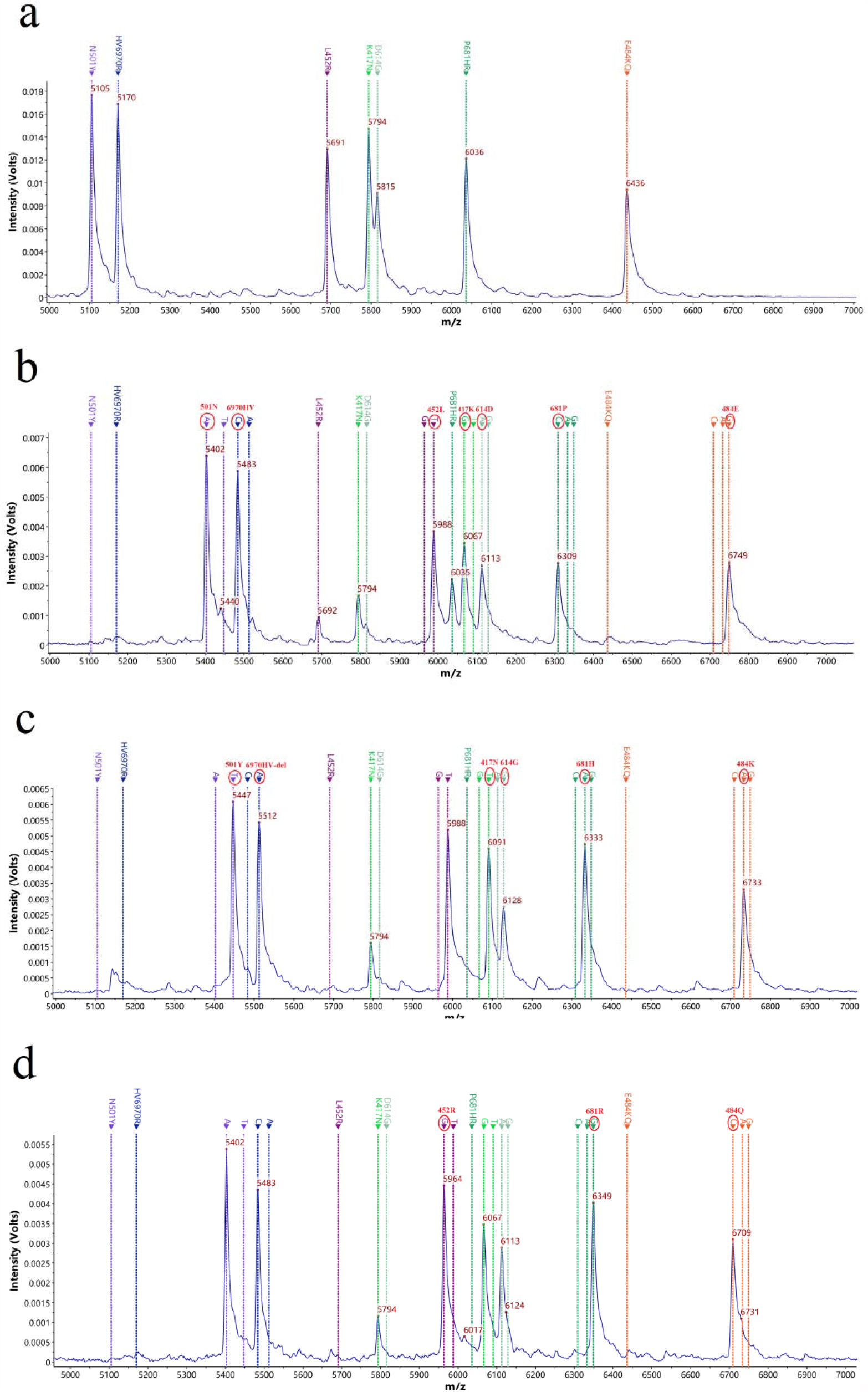
MS peak of the MPE probes. A: MS peaks of 9 MPE probes without extension; B: Target sites peaks of the MPE probes extended to non-SARS-CoV-2 variants; C: Target sites peaks of the MPE probes extended to SARS-CoV-2 S gene mutation plasmids 1(containing HV69-70del, K417N, E484K, N501Y, D614G, P681H mutation). D: Target sites peaks of the MPE probes extended to SARS-CoV-2 S gene mutation plasmids 2 (containing L452R, E484Q, P681R mutation).

### Detection limit and specificity of multiplex PCR-MALDI-TOF MS

The extended signals of mass probe pairs 5483±3/5512±3 (HV69-70del), 5402±3/5447±3 (N501Y), 6067±3/6091±3 (K417N), 6309±3/6333±3 (P681H), 6309±3/6349±3 (P681R), 6113±3/6129±3 (D614G), 6749±3/6733±3 (E484K), 6749±3/6709±3 (E484Q), and 5988±3/5964±3 (L452R) were not detected in 21 other respiratory pathogen nucleic acids and pharyngeal swab nucleic acids of healthy people, which showed that the specificity of this method was 100%. The detection limits for HV69-70del, K417N, E484K, N501Y, D614G, P681H, L452R, E484Q and P681R were 400, 1560, 400, 400, 400, 1560, 400, 400, and 1560 copies, respectively, and the total detection limit for all sites of the multiplex PCR-MALDI-TOF MS was 1560 copies when using the diluted concentration of the nucleic acids of the SARS-CoV-2 nonvariants (Fig. 2). The mutation sites were detected by synthetic DNA plasmids, and the detection limits of all the sites were between 100-400 copies.

**FIG 2.**
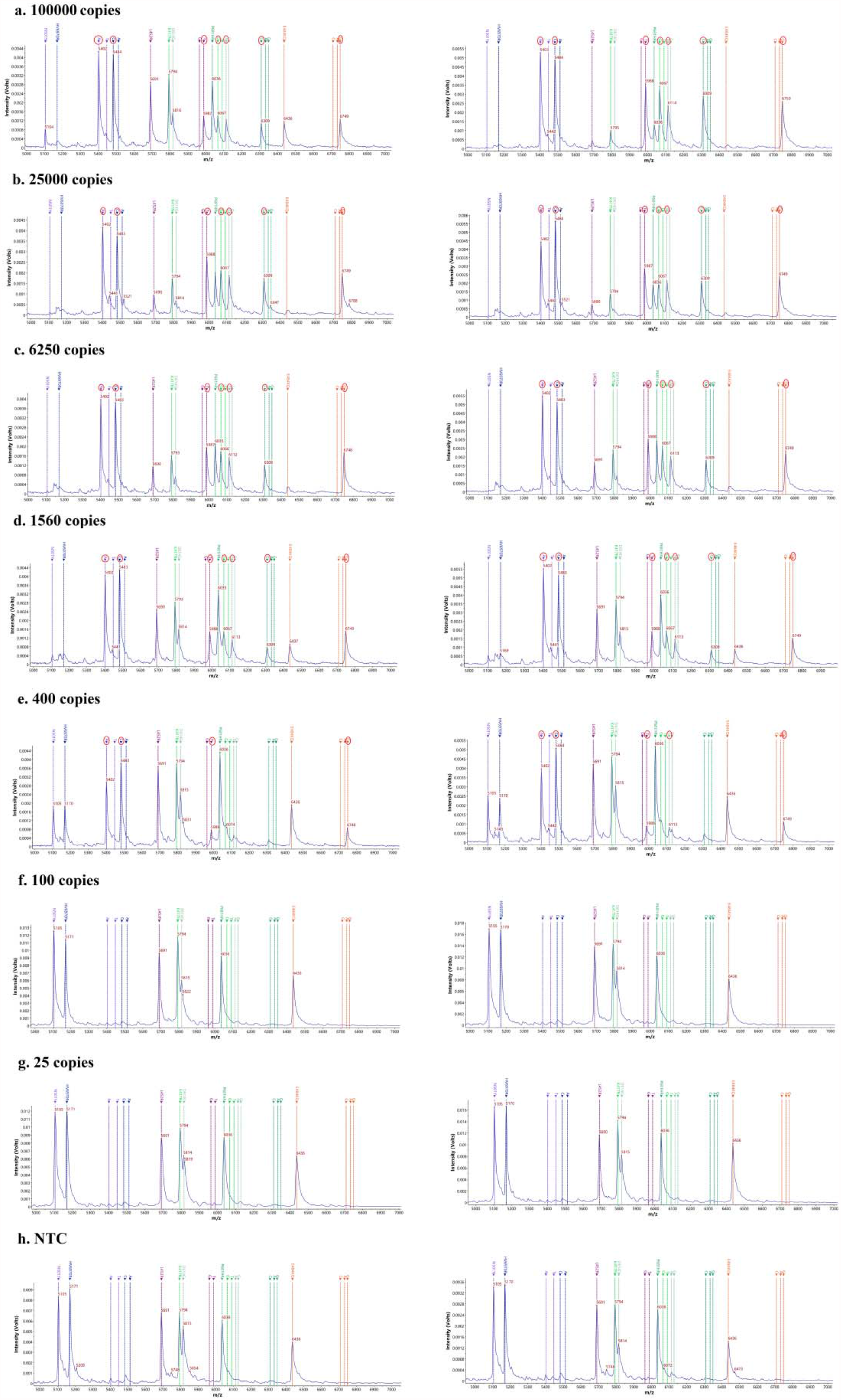
Detection limit of the 9 types of mutations at 7 mutated sites by non-SARS-CoV-2 variants. The left and right sides are two repetitions.

### Detection results of clinical nucleic acid specimens

The CT values of RT real-time PCR (FAM fluorescence signal CT value of N gene) of nucleic acid samples of 16 COVID-19 patients were 16.04-34.95, among which 10 samples with CT within 27 were all positive in the 7 target sites, and no mutation was found. Of the six specimens with CT values greater than 30, some of the sites were detected in 5, and none of the sites were detected in one specimen (Table 2).

**Table 2.** Detection results of target sites in COVID-19 patients by multiplex PCR-MALDI-TOF MS.

| Sample number | Specimen type | CT value | Target sites |  |  |  |  |  |  |
| --- | --- | --- | --- | --- | --- | --- | --- | --- | --- |
|  |  |  | N501Y | HV6970R | L452R | K417N | D614G | P681H/R | E484K/Q |
| 1 | sputum | 16.04 | A | C | T | G | A | C | G |
| 2 | sputum | 21.05 | A | C | T | G | A | C | G |
| 3 | sputum | 21.05 | A | C | T | G | A | C | G |
| 4 | sputum | 21.12 | A | C | T | G | A | C | G |
| 5 | sputum | 21.60 | A | C | T | G | A | C | G |
| 6 | sputum | 22.19 | A | C | T | G | A | C | G |
| 7 | sputum | 22.24 | A | C | T | G | A | C | G |
| 8 | sputum | 26.31 | A | C | T | G | A | C | G |
| 9 | sputum | 26.47 | A | C | T | G | A | C | G |
| 10 | sputum | 26.89 | A | C | T | G | A | C | G |
| 11 | sputum | 30.35 | NoCall | C | NoCall | NoCall | NoCall | NoCall | NoCall |
| 12 | BALF* | 30.89 | A | C | NoCall | NoCall | NoCall | NoCall | NoCall |
| 13 | sputum | 31.64 | A | C | NoCall | NoCall | NoCall | NoCall | NoCall |
| 14 | BALF | 32.34 | A | C | NoCall | NoCall | NoCall | NoCall | NoCall |
| 15 | sputum | 34.63 | A | C | NoCall | NoCall | NoCall | NoCall | NoCall |
| 16 | sputum | 34.95 | NoCall | NoCall | NoCall | NoCall | NoCall | NoCall | NoCall |

BALF: Bronchoalveolar lavage fluid.

## Discussion

In view of the emerging SARS-CoV-2 variants, researchers are trying to develop rapid detection methods in addition to genome sequencing. All six variants, B.1.1.7, B.1.351, B.1.429, B.1.526, P.1 and B.1.617, contain the D614G mutation, three contain the N501Y mutation ^3^, and the South African and Brazilian variants additionally contain mutations in E484K and K417N(T) ^3, 22^. The key SARS-CoV-2 mutations N501Y and D614G are the detection target sites that researchers mainly focus on. qPCR technology ^12^, RT-PCR technology, and CRISPR-Cas13a-based transcription amplification were used to detect the N501Y mutation site or D614G mutation site for the identification of SARS-CoV-2 variants ^13, 14^. Recently, a study highlighted that South African (K417N, E484K, N501Y) and Brazilian (K417T, E484K, N501Y)variants are more lethal than the UK variant (N501Y) ^5^. Based on the existing technology, all variants cannot be detected simultaneously by qPCR or CRISPR-Cas13a amplification technology. Herein, we developed a multiplex PCR-MALDI-TOF MS method that can simultaneously detect 9 mutation types in 7 mutated sites in the RBD of spike proteins HV69-70del (B.1.1.7),N501Y (B.1.1.7, B.1.351, P.1), K417N (B.1.315), P681H (B.1.1.7), D614G (B.1.1.7, B.1.351, P.1, B.1.429, B.1.526, B.1.617), E484K (B.1.351, P.1), L452R (B.1.617, B.1.429), P681R (B.1.617), and E484Q (B.1.617) to identify SARS-CoV-2 and its mutated variants. The workflow of this method is shown in Figure 4. Through combination of the results of the nine mutation sites, we can judge whether the detected sample is a SARS-CoV-2 infection, and if it is a SARS-CoV-2 infection, we can also determine which variant strain is present (Table 3).

**Table 3.** Interpretation basis of the identification of SARS-CoV-2 variants using multiplex PCR-MALDI-TOF MS.

| Mutation sites* | Identification sites and identification results (variants) |  |  |  |  |  |  |  |
| --- | --- | --- | --- | --- | --- | --- | --- | --- |
|  | India<br>(B.1.617) | UK<br>(B.1.1.7) | South<br>African<br>(B.1.351) | Clifornia<br>(B.1.429) | New York<br>(B.1.526) | Brazilian<br>(P.1) | Nonvariants | Non-SARS-CoV-2<br>infection |
| HV69-70R del | 5483 | ★5512 | 5483 | 5483 | 5483 | 5483 | 5483 | 5170 |
| L452R | 5964 | 5988 | ★5988 | ★5964 | 5988 | 5988 | 5988 | 5691 |
| N501Y | 5402 | 5447 | 5447 | 5402 | ☆5402 | ★5447 | 5402 | 5105 |
| D614G | 6113 | 6113 | 6113 | ★6113 | ★6113 | ★6113 | 6113 | 5816 |
| P681H | 6309 | ★6333 | 6309 | 6309 | 6309 | 6309 | 6309 | 6036 |
| P681R | ★6349 | 6309 | 6309 | ☆6309 | 6309 | 6309 | 6309 | 6036 |
| E484K | 6749 | 6749 | 6749 | 6749 | ★6733 | ★6733 | 6749 | 6436 |
| E484Q | ★6709 | 6749 | 6749 | ☆6749 | 6709 | 6749 | 6749 | 6436 |
| K417N | 6067 | 6067 | ★6091 | 6067 | ☆6067 | ☆6067 | 6067 | 5794 |

\*Mutation sites detected in this study. ★ and ☆ denote the mutation sites that must be identified and unidentified in the same test, respectively. The mass error of the spectrum peak value in this table is allowed to be less than 500 ppm ( $\pm 3$ Da).

India is suffering from a devastating wave of COVID-19. In the past few weeks, the number of cases and deaths has risen sharply. There are two “famous” mutations in the new variant B.1.617, which is widespread in India. One is L452R, which also exists in California variant. The other is E484Q, which is similar to the variant (E484K) first detected in South Africa and Brazil (Fig. 1D). A recently published study found that the L452R mutation may enhance the ability of SARS-CoV-2 to infect human cells in the laboratory. The California variant carries the same mutation, which increases the transmission ability by approximately 20% ^23^, and enhances the ability to escape the immune system, reducing the immune effect of the vaccine. If B.1.617 does increase the chance of re-infection (or breakthrough infection in vaccinated people), then this new variant may drive a surge in cases in other parts of the world. The method constructed in this study can simultaneously detect the two well-known mutation sites, L452R and E484Q, as well as P681R (similar to that of the South African variant, P681H in South Africa), which can accurately judge whether the sample is infected by Indian variant.

This method can not only detect SNP gene mutation, but also detect gene variation caused by deletion and insertion according to probe extension. The type of extended base was determined according to the peak value (m/z) acquired by MALDI-TOF MS. For example, for the HV69-70del site, if the peak of the detected extension product was 5483±3, then the extension base was C, indicating that catgtc (HV69-70R) was not deleted, and the detected sample was a non-SARS-CoV-2 variant; if the peak of the detected extension product was 5512±3, then the extension base was A, indicating that catgtc was deleted (Fig. 3), and the detected sample was a SARS-CoV-2 variant; if these two characteristic peaks were not detected, the sample was not infected with SARS-CoV-2. The detection of mutation sites through the same principle (Fig. 3).

**FIG 3.**
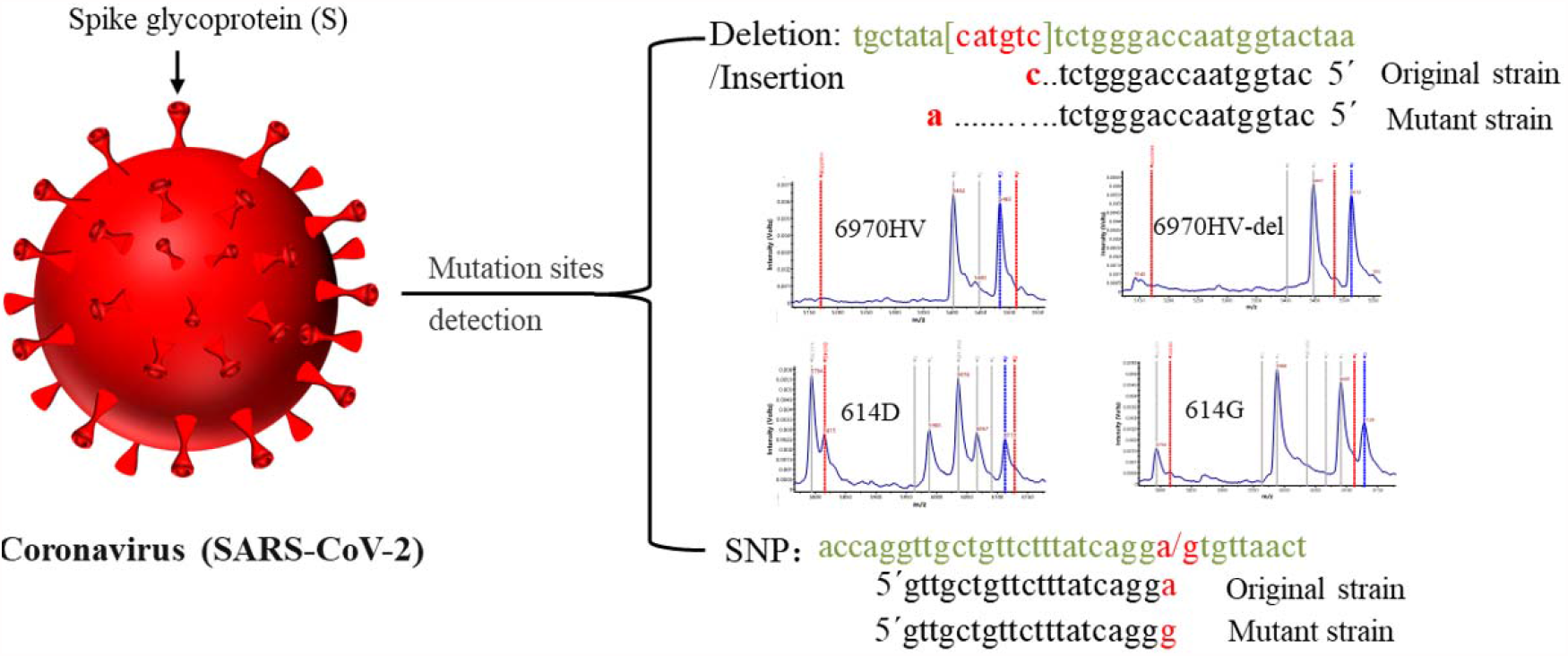
The detection principle of PCR-MALDI-TOF MS.

**FIG 4.**
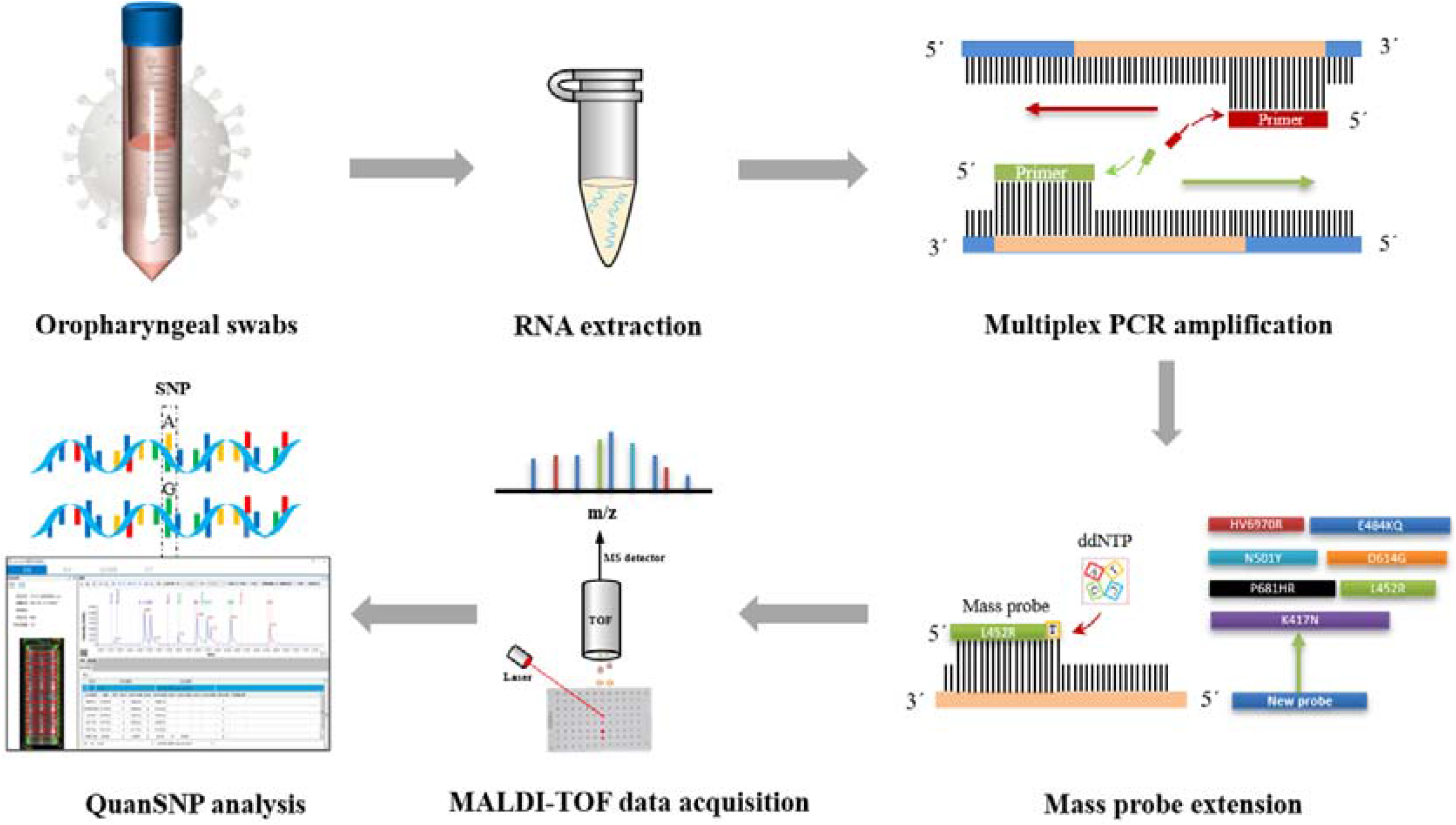
Workflow of multiplex PCR-MALDI-TOF MS for SARS-CoV-2 variant detection.

This method relies on MPE probe detection after PCR amplification and has high specificity. There was no nonspecific identification in various non-SARS-CoV-2 pathogens (9 bacteria, 11 respiratory viruses) or nucleic acid samples from non-COVID-19 patients. The detection method of mutation sites based on PCR and first generation sequencing technology has a high requirement on the quantity of PCR amplification products. PCR products with insignificant electrophoretic bands often fail to be sequenced due to insufficient quantity. However, the PCR-MALDI-TOF MS requires less PCR products than sequencing when extending MPE probe. The PCR products, which are not obvious in electrophoretic bands or even hard to be observed by naked eyes, can also be detected by MALDI-TOF MS after MPE extension. Therefore, for PCR-MALDI-TOF MS, the detection limit is affected by both multiplex PCR amplification and MPE probe extension, and there are great differences among different mutation sites. For example, D614G and P681H are located in the same PCR amplification fragment. The detection limit of D614G is 400 copies after MPE extension, while that of P681H/R is only 1560 copies, indicating that the probe extension efficiency of P681H/R is much lower than that of D614G, which is closely related to the base sequence near the detection site of P681H/R. Although the detection limit of the mutant site of synthetic plasmid is lower (100-400 copies), considering the lack of RNA virus reverse transcription step in the process of plasmid DNA amplification, we still use the LDL of nucleic acid amplification of SARS-CoV-2 infected samples (1560 copies) as the LDL of this method.

All 16 clinical samples were collected from February to March 2020, and no mutation sites were detected. The nucleic acid load of the samples with real-time PCR CT values greater than 30 was roughly estimated to be less than 1000 copies, which was lower than the detection limit of some sites. Therefore, only a portion of sites were detected in 5 samples with CT values greater than 30, and one sample was negative. The reliability of the method is verified by the detection of clinical samples, which indicates that the method has clinical application value.

PCR coupled with first generation sequencing technology is only suitable for the detection of multiple mutation sites in the nucleic acid sequence of less than 600 bp, and cannot be used for the detection of all mutation sites of SARS-CoV-2 (2000 bp). qPCR is only suitable for the detection of single mutation site. WGS is time-consuming, high cost and high technical requirements, so it is only suitable for mutation sites analysis in important samples, not suitable for routine sample screening. This method constructed in this study is very suitable for the detection of multiple mutation sites of SARS-CoV-2 variant. In addition, this method is open and extensible and can be used in a high-throughput manner (96 samples can be detected within 7 hours), easily allowing the addition of new mutation sites as needed to identify and track new SARS-CoV-2 variants as they emerge.

This study developed a novel strategy for detecting SARS-CoV-2 variants. However, this method has some limitations. First, the 3’ terminal sequence of MPE prove is fixed, and the quality of the probe is greatly influenced by the base sequence near the detection site, which can lead to the inefficient detection in extreme cases. Second, there is no SARS-CoV-2 variants in this study, and the detection limit of mutant plasmid can only be used as reference. Finally, the number of clinical specimens used in this study is limited, and the number of validation specimens needs to further increased.

As a simple screening assay for monitoring the emergence and spread of these SARS-CoV-2 variants, multiplex PCR-MALDI-TOF MS may be helpful for implementing public health strategies to counter these and future mutation strains.

## Data Availability

All the data in the manuscript met the specifications.

## Author contributions

Z.F. and X.D. conceived and managed the project, analyzed the data and wrote the manuscript. Z.J.Z. and L.T.Y. searched the literature. W.X.M. acquired the MS data. M.F.L and X.X.N. operated the experiments. H.X.X, Q.T. and Z.H.J. provide nucleic acids of clinical samples. L.J.X. and K.B. provided project related support.

## Reference

1. Tang, J.W., Tambyah, P.A. & Hui, D.S. Emergence of a new SARS-CoV-2 variant in the UK. J Infect 82, e27–e28 (2021).

2. Cascella, M., Rajnik, M., Aleem, A., Dulebohn, S.C. & Di Napoli, R. in StatPearls (Treasure Island (FL); 2021).

3. Galloway, S.E. et al. Emergence of SARS-CoV-2 B.1.1.7 Lineage - United States, December 29, 2020-January 12, 2021. MMWR Morb Mortal Wkly Rep 70, 95–99 (2021)

4. Zhou, D. et al. Evidence of escape of SARS-CoV-2 variant B.1.351 from natural and vaccine-induced sera. Cell 184, 2348–2361 e2346 (2021).

5. Khan, A. et al. Higher infectivity of the SARS-CoV-2 new variants is associated with K417N/T, E484K, and N501Y mutants: An insight from structural data. J Cell Physiol (2021).

6. Hou, Y.J. et al. SARS-CoV-2 D614G variant exhibits efficient replication ex vivo and transmission in vivo. Science 370, 1464–1468 (2020).

7. Volz, E. et al. Evaluating the Effects of SARS-CoV-2 Spike Mutation D614G on Transmissibility and Pathogenicity. Cell 184, 64–75 e11 (2021).

8. Laha, S. et al. Characterizations of SARS-CoV-2 mutational profile, spike protein stability and viral transmission. Infect Genet Evol 85, 104445 (2020).

9. Bull, R.A. et al. Analytical validity of nanopore sequencing for rapid SARS-CoV-2 genome analysis. Nat Commun 11, 6272 (2020).

10. Bal, A. et al. Two-step strategy for the identification of SARS-CoV-2 variant of concern 202012/01 and other variants with spike deletion H69-V70, France, August to December 2020. Euro Surveill 26 (2021).

11. Umair, M. et al. Whole-genome sequencing of SARS-CoV-2 reveals the detection of G614 variant in Pakistan. PLoS One 16, e0248371 (2021).

12. Durner, J. et al. Fast and cost-effective screening for SARS-CoV-2 variants in a routine diagnostic setting. Dent Mater 37, e95–e97 (2021).

13. Banada, P. et al. A Simple RT-PCR Melting temperature Assay to Rapidly Screen for Widely Circulating SARS-CoV-2 Variants. medRxiv (2021).

14. Wang, Y. et al. Detection of SARS-CoV-2 and Its Mutated Variants via CRISPR-Cas13-Based Transcription Amplification. Anal Chem 93, 3393–3402 (2021).

15. Jang, K.S. & Kim, Y.H. Rapid and robust MALDI-TOF MS techniques for microbial identification: a brief overview of their diverse applications. J Microbiol 56, 209–216 (2018).

16. Kostrzewa, M. Application of the MALDI Biotyper to clinical microbiology: progress and potential. Expert Rev Proteomics 15, 193–202 (2018).

17. Rahi, P., Prakash, O. & Shouche, Y.S. Matrix-Assisted Laser Desorption/Ionization Time-of-Flight Mass-Spectrometry (MALDI-TOF MS) Based Microbial Identifications: Challenges and Scopes for Microbial Ecologists. Front Microbiol 7, 1359 (2016).

18. Peng, J. et al. Type-specific detection of 30 oncogenic human papillomaviruses by genotyping both E6 and L1 genes. J Clin Microbiol 51, 402–408 (2013).

19. Peng, J. et al. Sensitive and rapid detection of viruses associated with hand foot and mouth disease using multiplexed MALDI-TOF analysis. J Clin Virol 56, 170–174 (2013).

20. Sjoholm, M.I., Dillner, J. & Carlson, J. Multiplex detection of human herpesviruses from archival specimens by using matrix-assisted laser desorption ionization-time of flight mass spectrometry. J Clin Microbiol 46, 540–545 (2008).

21. Zhao, F. et al. A multisite SNP genotyping and macrolide susceptibility gene method for Mycoplasma pneumoniae based on MALDI-TOF MS. iScience 24, 102447 (2021).

22. Nonaka, C.K.V. et al. Genomic Evidence of SARS-CoV-2 Reinfection Involving E484K Spike Mutation, Brazil. Emerg Infect Dis 27, 1522–1524 (2021).

23. Deng, X. et al. Transmission, infectivity, and antibody neutralization of an emerging SARS-CoV-2 variant in California carrying a L452R spike protein mutation. medRxiv (2021).

